# Alcohol Consumption and ALDH2 Polymorphism Jointly Modify the Association Between HDL Cholesterol and Coronary Artery Calcium Score in Older Adults

**DOI:** 10.64898/2026.01.30.26345249

**Authors:** Shao-Yuan Chuang, Ming-Ting Wu, Ren-Hua Chung, Chia-Hung Lai, Wan-Ju Cheng, Wen-Ling Liu, Chao Agnes Hsiung, Shu-Chun Chuang, I-Chien Wu, Chih-Cheng Hsu

## Abstract

**Importance:** The association between high-density lipoprotein cholesterol (HDL-C) and coronary artery calcium (CAC) burden remains unclear, and the potential modifying effects of lifestyle and genetic factors are not well characterized.

**Objective:** To clarify the relationship between HDL-C and CAC scores in older adults and examine whether alcohol consumption and the ALDH2 rs671 genetic variant modify this association.

**Design, Setting, and Participants:** Retrospective analysis of 819 community-dwelling older adults aged ≥55 years (378 men; 441 women) without a history of coronary stent placement, enrolled in the Healthy Aging Longitudinal Study in Taiwan. Baseline alcohol consumption and metabolic profiles were collected during 2014–2019; CAC scores were assessed in 2023 using low-dose computed tomography.

**Exposures:** Serum HDL-C levels, alcohol consumption status, and ALDH2 rs671 genotype.

**Main Outcomes and Measures:** CAC scores calculated using the Agatston method. Linear and logistic regression models assessed associations between HDL-C and CAC, adjusting for demographic and cardiometabolic risk factors. Interaction terms tested effect modification by alcohol consumption and ALDH2 genotype.

**Results:** HDL-C was inversely associated with log-transformed CAC scores after adjusting for age and sex (R = –0.228, *P* < .001). In multivariable models, older age, male sex, higher systolic blood pressure, elevated HbA1c, and lower HDL-C levels (β = –0.019, *P* = .003) were independently associated with higher CAC scores. The inverse association between HDL-C and CAC was significant among drinkers (β = –0.028, *P* = 0.005) and non-drinkers with ALDH2*GG* (β = –0.020, *P* = 0.0479) but not among non-drinkers without ALDH2*GG* (β = −0.005, *P* = 0.6106), with a significant interaction (*P* = 0.0276).

**Conclusions and Relevance:** Higher HDL-C levels were independently and inversely associated with CAC scores in older adults, with the relationship significantly modified by both alcohol consumption and ALDH2 genotype. These findings highlight a gene–environment interaction that may inform precision cardiovascular prevention strategies, particularly in populations with low prevalence of ALDH2 deficiency.

## Introduction

Cardiovascular disease (CVD) accounts for a significant proportion of deaths in the United States, globally, and within Asian populations. Large cohort studies have demonstrated that high-density lipoprotein cholesterol (HDL-C) is inversely associated with CVD risk in the U.S. population ^1,2^. Notably, this inverse association persists even among individuals with low-density lipoprotein cholesterol (LDL-C) levels below 70 mg/dL^3^. The protective effect of HDL-C against major cardiovascular events has been well established.

However, the relationship between HDL-C and coronary artery calcium (CAC) scores, a marker of subclinical coronary heart disease, remains inconsistent. The Dallas Heart Study reported no significant association between HDL-C and either prevalent coronary artery calcium or incident coronary heart disease (CHD) ^4^. Conversely, the Brazilian Longitudinal Study of Adult Health (ELSA-Brasil) indicated that lower HDL-C levels were positively associated with CAC incidence but not with CAC progression ^5^. In contrast, baseline HDL-C levels were inversely associated with CAC score progression in Asian populations ^6^. These findings underscore the complex and variable relationship between HDL-C and CAC scores, highlighting the need for further investigation.

The association between HDL-C and CAC scores may be confounded or modified by alcohol consumption. While moderate alcohol intake is typically associated with higher HDL-C levels^7^, heavy consumption can result in pathologically high HDL-C concentrations ^8^. This complexity extends to vascular health: light to moderate drinking is linked to lower CAC scores ^9,10^, whereas heavy consumption is positively associated with aortic calcification^11^. These findings suggest a complex interrelationship between alcohol, HDL-C, and arterial calcification. Furthermore, the impact of alcohol is significantly influenced by genetic variation in **Aldehyde Dehydrogenase 2 (ALDH2)** ^12^. The *ALDH2* rs671 polymorphism, which leads to enzyme deficiency, is a critical genetic marker associated with an increased risk of cardiovascular pathologies, including coronary artery disease, alcohol-induced cardiac dysfunction, heart failure, and drug-induced cardiotoxicity^13^. The complex relationship between HDL-C cholesterol and CAC score may be affected by alcohol consumption and **ALDH2 gene.**

Therefore, we hypothesize that (1) the association between HDL-C and CAC varies significantly based on the dose of alcohol consumption. In addition, (2) The ALDH2 rs671 deficiency—a key regulator of alcohol metabolism—acts as a critical genetic modifier that influences both HDL-C concentrations and CAC Score. By integrating genetic markers with lifestyle data, this study aims to clarify the divergent findings in existing literature and provide a more nuanced understanding of cardiovascular risk stratification in the context of alcohol-related metabolic pathways.

## Methods and Materials

### Study population

Healthy Aging Longitudinal Study in Taiwan (HALST) was founded by the National Health Research Institutes, Taiwan, and the baseline wave was initiated from 2008 to 2013. HALST is a prospective cohort study targeting investigating multidimensional determinants of healthy aging—including lifestyle behaviors and genetic, metabolic, and inflammatory factors—in an older Asian population. A total of 5663 community-dwellers aged 55 and over, recruited from seven selected cities/counties to represent the socio-demographic diversity of the Taiwanese population, participated in-home interviews and hospital-based clinical examinations in the first-wave survey (2009–2013), in the second-wave follow-up (2014–2019) and the third-wave follow-up is ongoing. The aims of the study and the collected information were reported in the prior study^14^ in detail.

For this study, older adults from the HALST cohort residing in Miaoli, Taoyuan, Changhua, and Kaohsiung were invited to undergo low-dose computed tomography (LDCT) scans to assess coronary artery health during 2023. As confirmed by physicians and medical records, 261 participants with coronary stents were excluded. Ultimately, 819 older adults (378 men and 441 women) without coronary stents were included in the study.

All study participants provided written informed consent before enrollment. The study protocol was reviewed and approved by the Institutional Review Board of the National Health Research Institutes, Taiwan (IRB No. EC1090903-R11). All procedures involving human participants were conducted in accordance with the ethical standards of the institutional and national research committees and with the 1964 Declaration of Helsinki and its later amendments.

### Study Design

This study was a retrospective design for investigating the association between baseline lipids and future coronary artery calcification scores (CAC Scores) in an older adult population. The baseline lipids were measured during 2014∼2019 and CAC scores were measured during 2023.

### Evaluation of coronary artery calcification score

LDCT was performed as the base of GOLD-CT (Geriatric Opportunity Low Dose CT) project. The GOLD-CT project was aimed to survey the lung nodule, emphysema, interstitial lung abnormality, coronary artery calcification, adipose tissue volume, muscle volume, bone density for opportunity quantitative assessments of multiple risk factors in the aging process. The CAC was assessed by a FDA-approved deep-learning based software (AView, CoreLine, South Korea) automatically and visually check by experienced readers.

The standard protocol of LDCT is 1. CTDI < 3mGy, total radiation dose < 1.5 mSVi for a 70 kG person. The scan range starts from thoracic inlet to the end of L2 vertebral body. Scan was performed during full inspiratory breath-hold (< 10 sec.) on a CT (>= 64 detector rows) scanner. Axial 1.0 mm slice thickness with standard kernal reconstruction were transferred to the central lab for analysis. The significance of CAC noted on LDCT, as compared to the standard full-dose ECG-gating CAC score, has been well validated^15–17^. The AI software automated CAC measurement has been validated in several studies^18^. The CAC scores were classified as zero, between zero and 10, between ten and 100, between 100 and 400, between 400 and 1000, and more than one thousand^19^. The Multi-Ethnic Study of Atherosclerosis ^20,21^ and systematic review ^22^showed that CAC score an dose-response relationship with incident atherosclerotic cardiovascular disease or coronary disease.

### Blood samples and the measurement of HDLC

All older adults were invited to participant the physical examinations in the hospitals. The brachial blood pressures were measured by Omron HEM 907 in a quiet room. All participants were measured the brachial systolic and diastolic blood pressure three times on right arm with appropriate cuff in the chair with back-support after five minutes resting. The average blood pressure from the second and third measures of blood pressure were used for this analysis. All older adults were asked to fast at least 8 hours for fasting blood biochemistries parameters, such as fasting glucose, triglycerides, high-density lipoprotein cholesterol (HDL-C), low-density lipoprotein cholesterol (LDL-Cholesterol), total cholesterol, and HbA1c. Smoking habits was classified into three groups, current smoker, never smoking and quite smoker, defined by the interviewed questionnaire. The medical histories of chronic diseases were defined by the face-to-face questionnaire and the status of taking medicine for the chronic diseases.

### Definitions of an alcohol drinker and alcohol consumption

Alcohol consumption habits were categorized as either current drinkers or non-current drinkers. Based on the total amount, frequency, and percentage of alcohol consumed per week, drinking levels were further classified into four categories: never, abstainers, moderate, and heavy drinkers.

Moderate drinking was defined as consuming no more than 14 units of alcohol per week for men and no more than 7 units per week for women, while heavy drinking was defined as consuming more than 14 units per week for men and more than 7 units per week for women. One unit of alcohol was equivalent to 10 grams of pure alcohol^23,24^. We classified those with quit of alcohol drinking into none-alcohol drinking, due to the retrospective design of this study.

### ALDH-2 polymorphic typing and classification

The ALDH2 polymorphism was identified using quality-controlled GWAS data ^25^. Specifically, the rs671 single-nucleotide polymorphism, a well-established functional variant of the ALDH2 gene, was selected and directly genotyped on the TWB2.0 array. This SNP was used to classify participants according to their ALDH2 genotype (AA, AG, or GG) to evaluate potential genetic modification of the association between HDL-cholesterol and coronary artery calcium score.

### Statistical methods

Continuous and categorical characteristics of the study population were summarized as mean (standard deviation [SD]) and number (percentage), respectively (Table 1). Coronary artery calcium (CAC) scores were summarized using the median and interquartile range (IQR) because of a skewed distribution. CAC severity was additionally categorized into five clinically relevant groups (0, 1–10, 11–100, 101–400, and ≥400) (Supplemental Table 1). Sex differences in CAC were assessed using appropriate non-parametric tests for continuous CAC and chi-square tests for categorical CAC groups; trends across ordered CAC categories were evaluated using the Cochran–Armitage trend test.

**Table 1.** Characteristics of study population.

|  | Mean | Standard deviation |
| --- | --- | --- |
| Age, yrs | 69.8 | 6.3 |
| Male gender, % | 46.15% |  |
| Body mass index, kg/m<sup>2</sup> | 24.4 | 3.4 |
| Waist, cm | 86.2 | 10.2 |
| Systolic BP, mmHg | 125.1 | 16.7 |
| Diastolic BP, mmHg | 68.9 | 9.3 |
| Pulse pressure, mmHg | 56.2 | 13.0 |
| Total cholesterol, mg/dL | 190.1 | 36.2 |
| LDL-cholesterol, mg/dL | 114.6 | 31.1 |
| HDL-cholesterol, mg/dL | 53.0 | 14.4 |
| Triglycerides, mg/dL | 121.2 | 78.1 |
| Fasting glucose, mg/dL | 106.5 | 23.0 |
| HBA1C, % | 5.98 | 0.82 |
| Uric acid, mg/dL | 5.8 | 1.39 |
| Hypertensive medicine, % (n) | 38.58% | (316) |
| Diabetes medicine, % (n) | 15.14% | (124) |
| Dyslipidemia medicine, % (n) | 17.09% | (140) |
| CAC score, median [p25, p75] | 112.6 | [23.17, 392.69] |
| Men | 178.2 | [41.5, 513.73] |
| Women | 83.9 | [15.22, 264.8] |
| Drink habits, % | 37.73% | (309) |
| Smoking, % | 9.52% | (78) |

For regression analyses treating CAC as a continuous outcome, CAC was log-transformed to improve normality. Because a subset of participants had very low CAC values (including zeros), a small constant was added prior to transformation (i.e., log[CAC + 0.1]) to accommodate zero values. Associations between lipid measures and metabolic factors and log-transformed CAC were initially explored in age- and sex-adjusted linear regression models (Table 2, Model 1). Multivariable linear regression was then used to identify factors independently associated with CAC by including clinically relevant covariates (age, sex, systolic blood pressure, body mass index [BMI], HbA1c, antihypertensive medication use, lipid-lowering therapy, and history of dyslipidemia) (Table 2, Model 2). Regression coefficients (β) with corresponding p-values were reported.

**Table 2.** Correlations of metabolic features at baseline for future CAC score (log-CAC)

|  | Univariate Analysis |  | Model 1 |  | Model 2† |  |  |
| --- | --- | --- | --- | --- | --- | --- | --- |
|  | Crude R | p-value | adjusted R* | p-value | Beta | 95% C.I. | p-value |
| Age, yrs | 0.166 | <.0001 |  |  | 0.036 | (0.014, 0.057) | 0.0012 |
| Male gender (M=1/W=2) | -0.180 | <.0001 |  |  | -0.661 | (-0.944, -0.377) | <.0001 |
| Body mass index, kg/m<sup>2</sup> | 0.215 | <.0001 | 0.215 | <.0001 | 0.067 | (0.025, 0.108) | 0.0017 |
| Waist, cm | 0.208 | <.0001 | 0.169 | <.0001 |  |  |  |
| Systolic BP, mmHg | 0.193 | <.0001 | 0.186 | <.0001 | 0.014 | (0.005, 0.022) | 0.0013 |
| Diastolic BP, mmHg | 0.053 | 0.127 | 0.080 | 0.0219 |  |  |  |
| Pulse pressure, mmHg | 0.209 | <.0001 | 0.196 | <.0001 |  |  |  |
| Total cholesterol, mg/dL | -0.128 | 0.0002 | -0.069 | 0.0495 |  |  |  |
| LDL-cholesterol, mg/dL | -0.045 | 0.2025 | -0.015 | 0.6725 |  |  |  |
| HDL-cholesterol, mg/dL | -0.274 | <.0001 | -0.228 | <.0001 | **-0.019** | **(-0.029, -0.009)** | **0.0003** |
| Triglycerides, mg/dL | 0.100 | 0.0044 | 0.125 | 0.0003 |  |  |  |
| Fasting glucose, mg/dL | 0.158 | <.0001 | 0.161 | <.0001 |  |  |  |
| HbA1C, % | 0.183 | <.0001 | 0.197 | <.0001 | 0.276 | (0.109, 0.443) | 0.0012 |
| Uric acid, mg/dL | 0.167 | <.0001 | 0.105 | 0.0027 |  |  |  |
| Hypertensive medicine, % | 0.253 | <.0001 | 0.241 | <.0001 | 0.509 | (0.209, 0.810) | 0.0009 |
| Diabetes medicine, % | 0.185 | <.0001 | 0.187 | <.0001 |  |  |  |
| Dyslipidemia medicine, % | 0.190 | <.0001 | 0.208 | <.0001 | 0.663 | (0.294, 1.031) | 0.0004 |
| Drink habits, % | -0.009 | 0.7957 | -0.056 | 0.1082 |  |  |  |
| Smoking, % | 0.070 | 0.0443 | 0.037 | 0.2858 |  |  |  |

\*Age and sex adjusted.
†: Multivariable linear regression with stepwise selection (R-square = 19.67%)
95% C.I. = 95% confidence intervals

To evaluate determinants of increasing CAC severity using categorical outcomes, multivariable ordinal logistic regression models were fitted with CAC category (0∼10, 11∼100, 101∼400 and 400+) as the dependent variable (Table 3). Odds ratios (ORs) and 95% confidence intervals (CIs) were estimated for higher CAC categories, adjusting for age, sex, systolic blood pressure, BMI, HbA1c, and use of antihypertensive and lipid-lowering medications.

**Table 3.** Determinants of CAC score in logistic regression.

|  | Odd ratio | Low of C.I. | Upper of C.I. | p-value |
| --- | --- | --- | --- | --- |
| Age, yrs | 1.039 | 1.018 | 1.061 | 0.0003 |
| Male gender (M=1/W=2) | 0.577 | 0.439 | 0.758 | <.0001 |
| Body mass index, kg/m<sup>2</sup> | 1.011 | 1.002 | 1.019 | 0.0116 |
| Systolic BP, mmHg | 1.052 | 1.011 | 1.095 | 0.0126 |
| HDL-cholesterol, mg/dL | 0.983 | 0.973 | 0.993 | 0.0008 |
| HBA1C, % | 1.290 | 1.096 | 1.520 | 0.0023 |
| Hypertensive medicine, % | 1.646 | 1.234 | 2.195 | 0.0007 |
| Dyslipidemia medicine, % | 2.189 | 1.527 | 3.138 | <.0001 |

The CAC-Score was classified into four groups: 1<sup>st</sup>: between zero and 10 (n=165, 20.5%), 2<sup>nd</sup>: between 11 and 100 (n=214, 26.13%), between 101 and 400 (n=239, 29.18%), and more than 400 (n=201, 24.54%).
The proportional odds testing: CHISQ= 12.29, degree of freedom=16, p-value=0.7238.

To assess potential non-linearity in the association between HDL cholesterol (HDL-C) and CAC, participants were categorized into sex-specific quartiles of HDL-C (<39, 39∼47, 47∼54 and 54+ for men; <47, 47∼56, 56∼66, and 66+ for women). Multivariable ordinal logistic regression models were fitted with HDL-C quartiles as the exposure, using the lowest quartile as the reference group (men: <39 mg/dL; women: <47 mg/dL). A p-value for trend was obtained by modeling quartile category as an ordinal term.

Effect modification by alcohol drinking or ALHD2 rs671 polymorphism was evaluated by including an interaction term between HDL-C and current alcohol drinking status (yes vs. no) or ALHD2 rs671 polymorphism (AA, AG, GG) in multivariable linear regression models of log-transformed CAC (Table 4), with adjustment for the covariates noted above.

**Table 4.** . The association between HDL-C and CAC score, stratified by alcohol habits, alcohol consumption and acetaldehyde dehydrogenase.

|  | Beta† (95% C.I.) | p-value |
| --- | --- | --- |
| **Model 1†** |  |  |
| Non-drinking habits (n=510) | -0.013 (-0.026, 0.001) | 0.0655 |
| Drinking habits (n=309) | -0.028 (-0.044, -0.013) | **0.0005** |
| p-value for interaction‡ |  | 0.0202 |
| **Model 2†** |  |  |
| ALDH2 |  |  |
| AA, (n=150) | -0.011 (-0.039, 0.017) | 0.4524 |
| GA, (n=270) | -0.017 (-0.036, 0.002) | 0.0786 |
| GG, (n=399) | -0.022 (-0.036, -0.008) | **0.0024** |
| p-value for trend |  |  |
| p-value for interaction‡ |  | 0.6899 |

†Models were adjusted age, sex, systolic blood pressure, body mass index, Hba1C, Hypertensive medicine, lipid lowering drugs.
‡The p-value for interaction was obtained from the interaction term in the multivariable model, which was adjusted for age, sex, systolic blood pressure, body mass index, HbA1c, antihypertensive medication use, and lipid-lowering drug use.

Stratified multivariable analyses were also conducted according to alcohol consumption (Yes vs. No) or ALHD2 rs671 polymorphism (AA, AG, GG) to estimate subgroup-specific HDL-C regression coefficients.

Furthermore, participants were cross-classified by alcohol drinking status and ALDH2 genotype to estimate subgroup-specific associations (Table 5). Three groups with combined alcohol drinker and ALDH2 genotype included (1) alcohol drinker, (2) non-alcohol drinker with ALDH2-GG genotype, and (3) non-alcohol drinker without ALDH2-GG genotype. The associations between HDL-C and CAC scores in the three groups were separately conducted in multivariable regression models with adjusted for age, sex, systolic blood pressure, body mass index, HbA1c, hypertensive medicine and lipid-lowering drugs. The interaction term of HDL-C x Drinking-Aldh2 (Three groups) was evaluated in the multivariable model. All statistical tests were two-sided, with p < 0.05 considered statistically significant. Analyses were performed using statistical software SAS 9.4.

**Table 5.** The association between HDL-Cholesterol and CAC Score in groups classified by drinking habits and aldehyde dehydrogenase deficiency Polymorphism.

| Drink status | ALHD2 | N | Beta | p-value | L-95 | U-95 |
| --- | --- | --- | --- | --- | --- | --- |
| No | Non-GG (AA/AG) | 286 | -0.005 | 0.6106 | -0.024 | 0.014 |
| No | GG | 224 | **-0.020** | **0.0479** | **-0.039** | **-0.000** |
| Yes |  | 309 | **-0.028** | **0.0005** | **-0.044** | **-0.012** |
| **p-value for interaction‡** |  |  | **0.0276** |  |  |  |

†: Models were adjusted for age, sex, systolic blood pressure, body mass index, HbA1C, Hypertensive medicine, and lipid-lowering drugs.
‡: Models were adjusted for age, sex, systolic blood pressure, body mass index, HbA1C, Hypertensive medicine, lipid-lowering drugs, and HDL-C x alcohol drinker\_ALDH2\_GG (Yes, No\_GG+, No\_GG-)(interaction term).

## Results

The 69.8 years of average age and 46.2% of male were in this population. The prevalence of hypertension, diabetes and dyslipidemia were 38.58%, 15.14% and 17.09%, respectively. There were 37.7% of current drinking and 9.52% of smoking.

### Descriptive Epidemiology of Coronary Artery Calcification Score

The median coronary artery calcium (CAC) score among all older adults was 112.6 (interquartile range [IQR]: 23.2–392.7). The distribution of CAC scores was as follows: 5.5% of participants had a CAC score of zero, 14.65% had scores between 1 and 10, 26.13% between 11 and 100, 29.18% between 101 and 400, and 25.54% had scores of 400 or greater (Supplemental Table 1).

Men exhibited significantly higher CAC scores than women, with median values of 178.2 and 83.9, respectively (p < 0.05). The corresponding proportions of CAC score categories for men and women were (1) CAC = 0: 2.91% (men) vs. 7.71% (women), (2) CAC 1–10: 13.49% vs. 15.65%, (3) CAC 11–100: 22.49% vs. 29.25%, (4) CAC 101–400: 31.75% vs. 27.89%, (5) CAC ≥ 400: 29.36% vs. 19.50%. These findings indicate a higher burden of subclinical atherosclerosis among men compared to women in this older adult population. (Supplemental Table-1)

### Determinants of CAC score

Older age, male sex, higher body mass index (BMI), larger waist circumference, elevated systolic blood pressure, and increased levels of fasting glucose and HbA1c were all positively associated with higher coronary artery calcium (CAC) scores. In age- and sex-adjusted analyses, HDL-cholesterol was significantly inversely associated with log-transformed CAC score (adjusted R = −0.228, *p* < 0.0001), whereas triglycerides were positively associated (adjusted R = 0.125, *p* < 0.0001). In contrast, LDL-cholesterol (adjusted R = −0.015, *p* = 0.6725) were not significantly associated with log-CAC (Table 2).

In multivariable linear regression models (Model 2, Table 2), systolic blood pressure, BMI, HbA1c, use of antihypertensive medications, and history of dyslipidemia were independently and positively associated with CAC score, while HDL-cholesterol remained significantly and inversely associated. Furthermore, multinomial logistic regression analysis (Table 3) revealed that older age, male sex, higher systolic blood pressure, elevated BMI and HbA1c levels, and use of antihypertensive or lipid-lowering medications were significantly associated with a greater CAC score category. HDL-cholesterol (odds ratio: 0.983; 95% confidence intervals: 0.973, 0.993), in contrast, was negatively associated with increasing CAC score categories.

We further categorized participants into four groups using sex-specific quartiles of HDL cholesterol to evaluate a potential non-linear association with CAC score in multivariable multinomial logistic regression. Using the lowest HDL-C quartile as the reference group (<39 mg/dL in men and <47 mg/dL in women), the odds ratios for higher (ordinal) CAC categories were 0.697 (95% CI, 0.472–1.029; p=0.0696) in the second quartile, 0.712 (95% CI, 0.474–1.071; p=0.1029) in the third quartile, and 0.543 (95% CI, 0.359–0.821) in the highest quartile. A significant inverse trend across increasing HDL-C quartiles was observed (p for trend=0.0073).

### HDL-C, CAC Score and alcohol drinking

The association between HDL-Cholesterol and CAC score was interacted with alcohol drinking. The negative association between HDL-cholesterol and CAC Score was stronger in alcohol drinker (beta = −0.028, p=0.0005), than those without alcohol drinking (beta = −0.013, p=0.0655) (p-value for interaction = 0.0202) in the multivariable with adjusted for covariates. (Table-4)

### Alcohol drinking and acetaldehyde dehydrogenase-2 polymorphism

Alcohol consumption was associated with polymorphism in the aldehyde dehydrogenase 2 (ALDH2, rs671) gene. The proportions of current alcohol drinkers across ALDH2 genotypes were 30.0% for AA, 33.0% for AG, and 43.9% for GG. A significant increasing trend in alcohol consumption with the presence of G alleles was observed (*p*-value for trend <0.001) (Supplemental Table 4).

### HDL-C, CAC Score, and ALDH2 Polymorphism

In stratified multivariable models adjusted for age, sex, systolic blood pressure, BMI, HbA1c, antihypertensive medication use, and lipid-lowering therapy, the association between HDL-C and CAC score also varied by ALDH2 genotype.

The beta of HDL-C for CAC score was −0.011 (*p* = 0.4524) for AA genotype, was −0.017 (*p* = 0.0786) for AG genotype, was −0.022 (*p* = 0.0024) for GG genotype. These findings suggest that both alcohol consumption and ALDH2 genetic variation may modify the relationship between HDL-C and subclinical atherosclerosis (Table 4).

The association between high-density lipoprotein cholesterol (HDL-C) and coronary artery calcium (CAC) score was examined in groups stratified by combined alcohol consumption and ALDH2 rs671 polymorphism status.

A significant inverse association between HDL-C and CAC scores was observed in the following subgroups: alcohol drinkers (β = −0.028, *p* = 0.0005) and non-drinkers with the GG genotype (β = −0.020, *p* = 0.049), but not in the non-drinker without GG genotype (β = −0.005, *p* = 0.6106). The interaction between alcohol tendency and HDL-C was broadline statistically significant (*p* = **0.0276** for interaction). (Table-5)

## Discussion

Lower HDL-cholesterol (HDL-C) levels were associated with higher subsequent coronary artery calcium (CAC) scores in this retrospective cohort of older adults. Effect modification by alcohol use was evident: an inverse association between HDL-C and CAC was observed among drinkers, but not among non-drinkers (p for interaction = 0.0202; Table 4). In addition, rs671 genotype was strongly related to alcohol consumption (p for trend < 0.0001; Supplemental Table 4). The HDL-C–CAC association was significant in participants with the rs671GG genotype (β = −0.022, p = 0.0024). In stratified analyses incorporating both alcohol use and rs671, an inverse association persisted among drinkers (β = −0.028, p=0.005) and among non-drinkers who carried rs671GG (β = −0.020, p = 0.0479), but not among non-drinkers without rs671GG (β = −0.005, p = 0.6105), with a significant interaction (p = 0.0276). Collectively, these results indicate that alcohol consumption and ALDH2 rs671 jointly modify the relationship between HDL-C and CAC.

The mechanism underlying the association between moderate alcohol consumption and elevated HDL-cholesterol levels involves several biological pathways. First, ethanol may enhance hepatic expression of apolipoprotein A-I (ApoA-I), a key structural protein of HDL, thereby promoting HDL particle formation^26^ Second, HDL cholesterol facilitates reverse cholesterol transport (RCT), the process of transporting excess cholesterol from peripheral tissues to the liver. Alcohol consumption has been shown to increase the activity of key enzymes involved in RCT, including lecithin-cholesterol acyltransferase (LCAT)^27^ and cholesteryl ester transfer protein (CETP) ^28^. Third, polyphenols such as resveratrol in red wine may exert additive effects by further enhancing HDL-C levels and function beyond those attributable to ethanol alone^29^. Most of the alcohol drinkers in this study were moderate consumption level, and those had elevated age-and sex-adjusted HDL-C levels. ALDH2 polymorphism was associated with alcohol behaviors in studies among the Asian population.

The ALDH2 genotype is one of the most robust genetic determinants of alcohol consumption patterns in Asian populations^30^. A genome-wide association study identified **rs671** as the top variant associated with drinking behavior in Japanese populations^31^. The ALDH2 genotype influences the level of alcohol consumption, with a clear inverse linear relationship observed between the *Lys* (*2) allele and alcohol intake in Japanese men^32^. Moreover, a similar association between ALDH2 genotype and alcohol consumption has been reported in the Korean population^33^. Our findings are also consistent with these prior studies, demonstrating that the proportion of current alcohol drinkers was positively and linearly associated with the frequency of the *G* allele.

The association between HDL-Cholesterol and CAC score remains ambiguous. The Dallas Heart Study reported no significant association between HDL-C and either prevalent coronary artery calcium or incident coronary heart disease (CHD) ^4^. Moreover, A systematic review and meta-analysis with five prospective cohort studies revealed no significant protective effect of high HDL-C against CAC >0^34^. Conversely, the Brazilian Longitudinal Study of Adult Health (ELSA-Brasil) indicated that lower HDL-C levels were positively associated with CAC incidence but not with CAC progression ^5^. In contrast, baseline HDL-C levels were inversely associated with CAC score progression in Asian populations ^6^. Our study supports a significant inverse association between HDL-C and coronary artery calcium (CAC) score. Additionally, we found that alcohol consumption may modify this relationship, with a stronger inverse association observed among individuals who consumed alcohol compared to those who did not (p-value for interaction = 0.0202). The degree of alcohol intake further influenced the association: the strongest inverse relationship was found in heavy drinkers, followed by moderate drinkers, non-drinkers, and those who had quit drinking, who showed the weakest association. Furthermore, the interaction between alcohol consumption and aldehyde dehydrogenase 2 (ALDH2) polymorphism slightly modified the association. Additionally, ALDH2 polymorphism combined with alcohol consumption slightly modified the association (p-value for trend = 0.0276), and the alcohol drinkers had the strongest negative association between HDL-C and CAC Score (beta = −0.028, p=0.0005), and the non-drinker with ALDH2-GG had a modest association (beta = −0.020, p=0.0479). These findings could provide a possible explanation for the inconsistencies found in prior studies.

Although higher HDL-cholesterol (HDL-C) has historically been inversely associated with coronary artery disease (CAD) in observational cohorts, multiple lines of evidence indicate that HDL-C concentration is not uniformly cardioprotective. Mendelian randomization analyses suggest that genetically elevated HDL-C does not necessarily reduce myocardial infarction risk (PMID: 22607825). In parallel, pharmacologic approaches that raise HDL-C have largely failed to reduce cardiovascular events, including niacin added to intensive statin therapy^35^ and extended-release niacin/laropiprant in high-risk patients^36^, while some CETP-inhibitor strategies were harmful^37^.

These findings support the concept that HDL “quantity” may not capture HDL “quality,” and that HDL function (e.g., cholesterol efflux capacity) may be more relevant to atheroprotection than HDL-C concentration alone^38,39^. Coronary artery calcium (CAC), a robust marker of coronary atherosclerotic burden, shows a clear dose–response relationship with incident ASCVD in large multiethnic cohorts and meta-analytic evidence^20,22,21^. Within this context, our data demonstrated that elevated HDL-C was significantly inversely associated with CAC score, but this association was modified by alcohol consumption and ALDH2 rs671. Specifically, the inverse association between HDL-C and CAC score was evident among current alcohol drinkers, but not among non-drinkers, and was present among non-drinkers with the rs671-GG genotype, but not among non-drinkers without the rs671-GG genotype. Alcohol intake is known to increase HDL-C in intervention studies^40^ and has been associated with CAC prevalence/incidence/progression in MESA^41^, providing a plausible exposure pathway for effect modification. Additionally, ALDH2 rs671—a functional East Asian variant influencing acetaldehyde metabolism and drinking behavior—has been associated with coronary heart disease susceptibility in Asian populations^42^, supporting the interpretation that gene–environment interactions may shape how HDL-C relates to subclinical coronary atherosclerosis.

Several limitations of this study should be acknowledged. First, because of the retrospective design, baseline CAC scores were unavailable; therefore, we could not evaluate the longitudinal association between baseline HDL-C levels and subsequent CAC progression. Pre-existing, unmeasured CAC at baseline could have biased the observed association in either direction, depending on its distribution across HDL-C strata. If individuals with higher HDL-C already had lower CAC at baseline, the absence of baseline adjustment would tend to attenuate the inverse association observed at follow-up. Accordingly, the association between baseline HDL-C and later CAC in our analysis is likely to be underestimated. Second, alcohol consumption was assessed using a structured questionnaire, which may be subject to recall or reporting bias. Third, the study population consisted exclusively of Han Chinese individuals; thus, the findings may no t be generalizable to populations with different genetic backgrounds, particularly those in which the ALDH2 polymorphism is rare (e.g., Caucasian populations).

Despite these limitations, the study has several strengths. CAC scores were objectively assessed using low-dose CT scanning. The observed associations between alcohol consumption, ALDH2 polymorphism, and HDL-C levels are consistent with previous findings^43,44^, providing external validation. Notably, similar prospective studies such as MESA^43^ and the South Bay Heart Watch^44^ have examined comparable associations. Additionally, we adjusted for a range of potential confounders, including alcohol consumption, in multivariable models, and demonstrated a robust inverse association between HDL-C and CAC scores. Importantly, our findings highlight a significant interaction between HDL-C and both alcohol consumption and ALDH2 genetic variation, suggesting that alcohol-related behavior and genetic predisposition may modify the relationship between HDL-C and subclinical atherosclerosis.

## Conclusion

In this cohort of older adults without known coronary stents, HDL-cholesterol was independently and inversely associated with coronary artery calcium (CAC) scores. The inverse association was particularly evident among participants with alcohol drinking habits, and a significant interaction with alcohol use or carrying the rs671*GG was observed. These findings suggest that HDL-C may exert a protective effect against subclinical coronary atherosclerosis and underscore the importance of accounting for gene–lifestyle interactions in cardiovascular risk stratification.

## Results

## Data Availability

The data that support the findings of this study are available from the corresponding author upon reasonable request. Access to the underlying participant-level data is subject to approval by the study?s research committee and institutional governance requirements. Data will be shared through a research collaboration framework, including review of the study proposal and execution of an appropriate data use agreement to ensure participant privacy and compliance with ethical and legal regulations.

---

**Supplemental Table 1.**
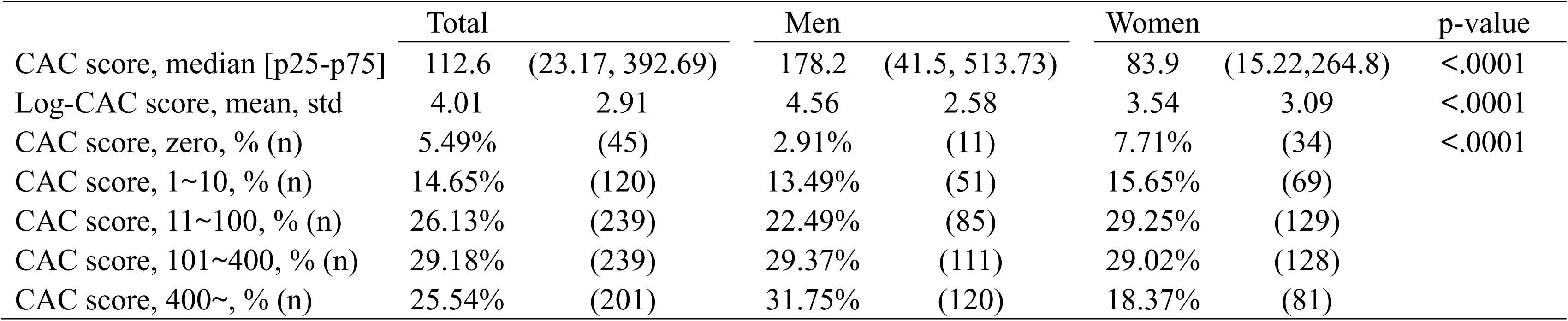
The distribution of the coronary artery calcium score of the population.

**Supplemental Table 2.** The association between alcohol consumption and CAC-Score.

|  |  | CAC-score |  | Log-CAC |  |
| --- | --- | --- | --- | --- | --- |
| Total | n | Mean±sd | Median [q1, q3] | Mean† ± s.e | p-value |
| Quit-3 | 86 | 327.7±509.4 | 152.0 [28.9, 444.5] | 4.26 ± 0.23 | 0.9117 |
| None-2 | 424 | 335.3±646.5 | 122.6 [23.3, 367.2] | 4.52 ± 0.13 | 0.0633 |
| Moderate-0 | 263 | 294.6±496.3 | 102.6 [17.3, 346.2] | 4.09 ± 0.13 | Reference |
| Heavy-1 | 46 | 379.6±454.7 | 215.2 [33.4, 511.1] | 4.47 ± 0.31 | 0.6514 |
| Men | n | Mean±sd | Median [q1, q3] | Mean† ± s.e | p-value |
| Quit-3 | 60 | 344.8±579.2 | 164.4 [31.2, 399.1] | 4.53 ± 0.26 | 0.8485 |
| None-2 | 109 | 483.1±789.5 | 205.8 [56.2, 615.2] | 4.86 ± 0.19 | 0.9891 |
| Moderate-0 | 168 | 397.7±585.3 | 164.3 [41.6, 511.7] | 4.78 ± 0.16 | Reference |
| Heavy-1 | 41 | 409.3±473.0 | 337.9 [33.4, 575.3] | 4.86 ± 0.32 | 0.9953 |
| women | n | Mean±sd | Median [q1, q3] | Mean† ± s.e | p-value |
| Quit-3 | 26 | 288.1±297.2 | 100.4 [24.0, 547.2] | 4.42 ± 0.40 | 0.0816 |
| None-2 | 315 | 284.1±581.8 | 97.8 [18.6, 276.4] | 4.16 ± 0.12 | 0.0036 |
| Moderate-0 | 95 | 112.2±159.1 | 40.3 [1.7, 163.0] | 3.34 ± .021 | Reference |
| Heavy-1 | 5 | 136.3±79.4 | 134.8 [126.6, 199.3] | 4.60 ± 0.92 | 0.5366 |

†: adjusted for age and sex

**Supplemental Table 3.**
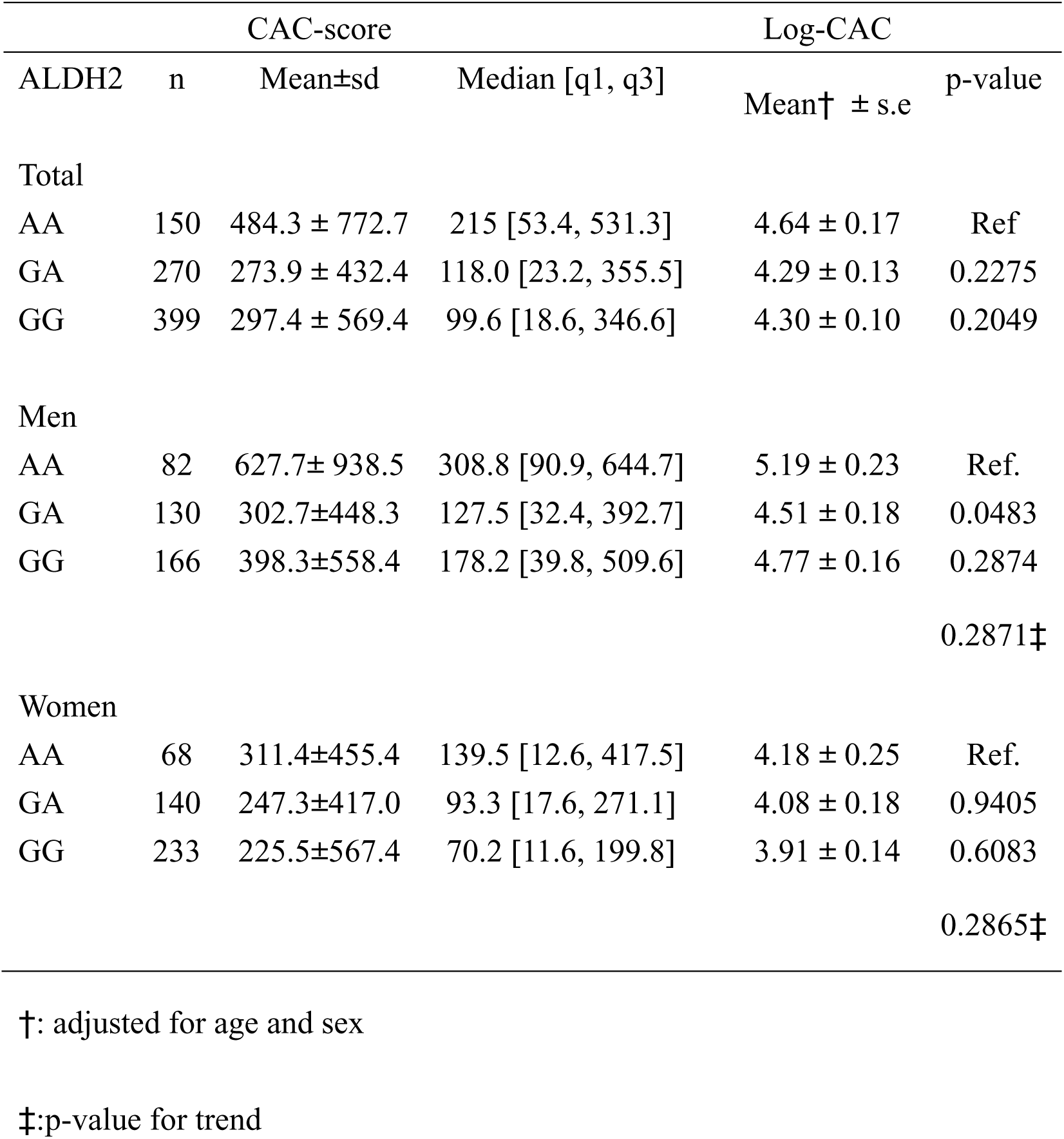
The association between ALDH2 and CAC-Score.

**Supplemental Table 4.** The association between ADLH2 and alcohol consumption.

| Alcohol consumption | ALDH2 |  |  | Total |
| --- | --- | --- | --- | --- |
|  | AA | AG | GG |  |
| Quit-3 | 18(12.0%†) | 21 (7.8%) | 47 (11.8%)) | 86 (10.5%) |
| None-2 | 87 (58.0%) | 160 (59.3%) | 177 (44.4%) | 424 (51.77%) |
| Moderate-0 | 38 (25.3%) | 81 (30.0%) | 144 (36.1%) | 263 (32.11%) |
| Heavy-1 | 7 (4.7%) | 8 (3.0%) | 31 (7.8%) | 46 (5.6%) |
| Non-drinking | 70.0% | 67.0% | 56.1% | 61.28% |
| drinking | 30.0% | 33.0% | 43.9% | 38.72% |
| Total | 150 (18.3%‡) | 270 (32.97%) | 399 (48.72%) | 819 |

†: Column percentage
‡: Row percentage
p-value for trend < 0.001
The relationship between alcohol consumption and ALDH2 rs671 genotype (AA, AG, GG) was assessed using chi-square tests, with a test for trend performed across genotypes by the number of G alleles (Supplemental Table 4).

**Figure 1:**
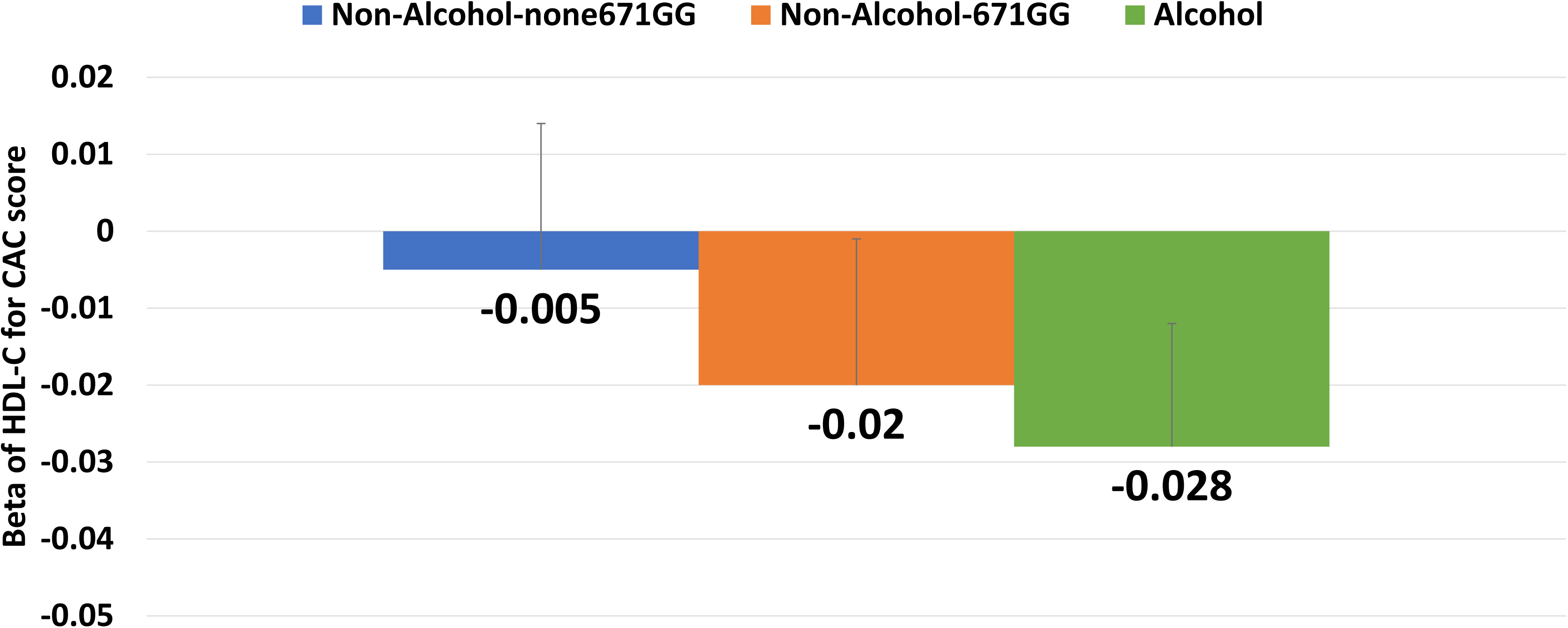
Associations of HDL-Cholesterol and Coronary Artery Calcium Score, stratified by drinking and ALDH2
Models were adjusted for age, sex, systolic BP, body mass index, medications for hypertension, diabetes and dyslipidemia, and
smoking. *Adjusted mean ± standard error.

